# Current Practices in Stroke Systems of Care: An International Survey of Progressive Stroke System Processes

**DOI:** 10.1101/2022.10.10.22280927

**Authors:** Matthew E Ehrlich, Shreyansh Shah, Brad J Kolls, Mayme Roettig, Lisa Monk, Carmelo Graffagnino

## Abstract

**Background and Aims:** Guidelines by the AHA/ASA recommend the development of stroke systems of care yet do not provide specifics as to how this should be done. As a first step in developing a regional stroke system of care in the “Stroke Belt” we sought to understand how high performing stroke systems across the world were organized in order to identify best practices and opportunities for meaningful system improvement.

**Methods:** An 81-question survey was developed to examine current practices in high-performing stroke systems. Twenty stroke centers worldwide were invited to participate. The survey encompassed all aspects of stroke systems of care from acute care and EMS practices to IT and discharge. Data analysis was conducted using REDCap internal analytic tools.

**Results:** Nine of 20 invited centers (45%) completed the questionnaire. Responding centers reported annual averages (median) of 750 ischemic strokes (range 350-1444) and 125 (100-501) hemorrhagic strokes, 150 (60-440) IV alteplase administrations, and 55 (12-130) endovascular thrombectomy procedures. At all 9 systems, EMS providers are trained in identifying stroke and utilize prehosptial stroke scales. Six (66.7%) reported destination protocols based on stroke severity involving bypass to endovascular thrombectomy capable (EVT-C) centers. All centers report expedited referral hospital transfer processes for neurointervention. Five (55.6%) centers reported patients with confirmed large vessel occlusions bypass the emergency department and are transported directly to the neurointerventional suite.

**Conclusions:** This international survey reveals useful practice patterns and processes used by progressive stroke systems. This information will be integrated into our regional system of care quality improvement program.

## Introduction

Stroke remains the 5^th^ leading cause of death in the United States, with increasing prevalence, and 2^nd^ leading cause of death globally.^1,2^ Recent Centers for Disease Control (CDC) data also indicates that despite nearly four decades of decline in stroke death rates, this decline has begun to stagnate and even reverse in many “Stroke Belt” states in the Southeast US.^2,3^ Further, recent advances in endovascular therapy (EVT) for acute ischemic stroke caused by large vessel occlusion (LVO)^4–10^ have magnified the need for rapid access to advanced specialty providers and therapeutics, which are not widely available. In light of this rapid paradigm shift in acute stroke care, health systems must quickly evolve. This will require large-scale regionalization approaches. Recently published American Heart Association/American Stroke Association (AHA/ASA) guidelines for the treatment of acute ischemic stroke recognized this with a high level recommendation (1A) that “regional systems of stroke care should be developed”.^11^

In response to these needs, especially in a region of the United States with the highest rates of stroke mortality^3^, we developed a comprehensive regional approach to improve stroke care through the application of best clinical practices utilizing a systems of care model quality improvement program known as Implementation of Best Practices for Acute Stroke care – Developing and optimizing regional systems in stroke Care (IMPROVE Stroke Care). IMPROVE Stroke Care utilizes implementation science approaches that have previously been demonstrated to be highly effective in other time-sensitive vascular emergencies such as ST elevation myocardial infarction (STEMI) and cardiac arrest.^12–14^ The primary vision for the IMPROVE Stroke Care project is to reduce disability and mortality from acute stroke in the region by identifying optimal processes of stroke care delivery through sharing data on performance and best practice, allowing redesign of systems within the partnered networks, leveraging existing local hub-and-spoke model hospital systems. The methods for this program have been previously published.^15^

## Aims

The first phase in the development of the IMPROVE Stroke Care program was to evaluate the current state of advanced stroke systems worldwide, and identify best practice processes to inform and provide evidence for recommendations included within the program’s Manual of Operations. High-level clinical guideline statements exist from organizations such as the AHA/ASA and the European Stroke Organization (ESO). Additional practice guidance can be found in large clinical trials, and big-data reviews from large data repositories such as Get With The Guidelines – Stroke. However, cutting-edge approaches are often not available from these sources, simply because they are too new to have big-data backing, or large clinical trials to support it, which take considerable time and funding. Additionally, expert consensus opinion level data on current practice controversies and innovative science is often lacking in published literature, or difficult to locate and summarize. We developed this international survey of high-performing and progressive stroke centers to obtain this data and inform advanced practice recommendations for the IMPROVE Stroke Care Program Manual of Operations.

## Methods

An 81-question survey was developed with input from team members with backgrounds in stroke, neurocritical care, emergency medicine, health services research, quality improvement and nursing. Questions in the survey tool included center demographics, patient volumes, staffing protocols, acute stroke code processes, Endovascular therapy (EVT) processes, Emergency Medical Systems (EMS), stroke IT and data collection, telestroke use, patient transfer processes, post-acute patient management and system feedback methodology. Data was collected from January 2017 to May 2017. The entire survey tool can be found in appendix 1. Twenty advanced stroke systems worldwide were identified based on publications, conference presentations from investigators at these institutions, as well as colleague input, representing a convenience sample. Physician leaders of their respective stroke programs at those institutions were contacted via email and invited to participate in the study. The survey and database were built, and study data were collected and managed, using RedCap^16^ electronic data capture tools hosted at Duke University. Statistical analysis was conducted using RedCap internal statistical tools. Basic counts and percentages, averages (mean or median as appropriate) and range were calculated as appropriate for individual survey element responses.

All study procedures were deemed exempt from review by the local Institutional Review Board. IMPROVE Stroke Care is funded by grants from the Medtronic Foundation, Daiichi Sankyo, and Chiesi. The funding sources had no involvement in the design or conduct of the study, data management or analysis, manuscript preparation or authorization for submission.

## Results

### Center demographics

Of 20 invited stroke centers, 9 responded with complete surveys (45%) (Figure 1). Seven of these centers self-identified as “Comprehensive Stroke Center” as defined by the US Joint Commission of Hospital Certification, or equivalent certifying agencies in Canada and Europe; 2 identified as “Intervention capable”, defined as primary stroke center with endovascular capability but without the official credentials of “comprehensive stroke center”. The survey pre-dated official separate certification by The Joint Commission for Thrombectomy-Capable Stroke Centers. On average (median) the 9 responding centers cared for 750 ischemic stroke patients per year (range 350-1444) and 125 primary intracerebral hemorrhagic strokes per year (range 100-501); in total, 81.4% percent ischemic and 18.6% primary intracerebral hemorrhage. Additionally, these centers reported a median 80 (range 47-177) primary subarachnoid hemorrhages (SAH) cared for yearly. Participating centers reported a median of 150 acute ischemic stroke patients treated with IV alteplase (tissue plasminogen activator -tPA) (range 60-440) and an average of 86 endovascular thrombectomy procedures per year (12-130).

**Figure 1.**
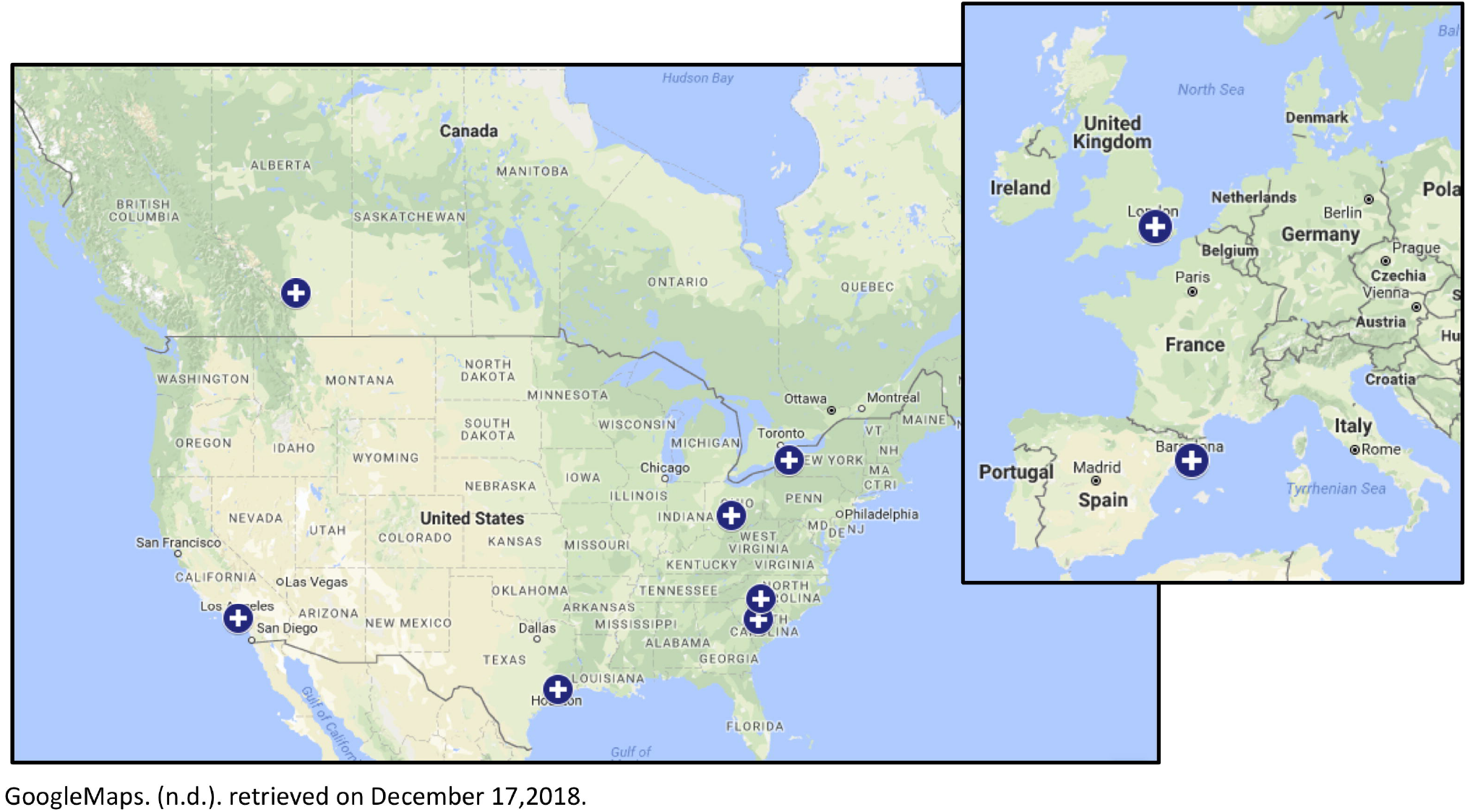
Map of participating centers.

### Staffing

Participating centers employ an average of 5 vascular neurologists (3-12) and 3 vascular neurosurgeons (1-5) on faculty, as well as 2 neurointerventionalists (from either radiology, neurosurgery or neurology departments) (2-7). There is a neurologist presence on site 24/7 in 7 (77.8%) of centers, and 77.8% also report having a dedicated Stroke/Neurovascular service in the hospital. Eight of the nine centers’ neurointerventional suites are open 24/7/365, however only 3 (33.3%) report that a neurointerventionalist responds to all stroke codes to evaluate eligibility for EVT. All but one center have a ‘back-up interventionalist’ available in case of multiple cases needing simultaneous intervention, and 6 (66.7%) have a nearby second intervention-capable center they defer or transfer to if their own suite is unavailable. Of the 9 responding centers, 8 (88.9%) have a dedicated Neurocritical Care Unit, and employ an average of 2 (0-9) specialty trained neuro-intensivists to staff them.

### EMS Services

When queried regarding supporting EMS agency stroke training, 8 centers report their local EMS responders in all EMS vehicles have training in identifying patients at risk for stroke based on chief complaint or presenting symptoms, while the remaining center noted EMS personnel in at least >50% of their vehicles had such training. All 9 hospital systems report these trained EMS personnel utilize a pre-hospital stroke scale to evaluate these patients; scales used included Cincinnati prehospital stroke scale (4), a modified version of Cincinnati (2), LAMS (1), MEND (1), RACE (2), FAST(1), LAPSS (2) (a single center could report multiple scales used due to multiple EMS agencies). EMS destination protocols based on stroke severity was reported by 6 centers (66.7%), with this same number reporting leadership had worked closely with their EMS agencies to develop these destination protocols. Though the criteria for initiating bypass to an interventional-capable hospital varied by center, common responses included utilizing severity scores (RACE >4 or LAMS ≥4); at least two centers commented on distance to CSC in addition to severity scores (“if CSC within 30minutes” plus LAMS ≥4). Mobile Stroke Units (MSU) were used by only 2 of the 9 centers (22.2%). None of the participating systems reported their EMS services to be utilizing commercial technologies or mobile applications to track stroke patients and processes in real-time. When EMS stroke assessment tools identify potential stroke patients, 4 (44.4%) of centers have patients bypass ED evaluation to go directly to imaging (CT/MRI) on arrival. In these 9 centers, an average of only 56.9% (33%-83%)of patients are brought via EMS.

### Acute Therapies and management

Within the ED, 66.7% of centers allow nursing staff to activate acute stroke codes. Every center noted a staff neurologist makes the final decision regarding administration of alteplase in acute stroke cases. Only 2 sites indicated utilizing telestroke services internally in some capacity, and only a single center indicated leveraging their own providers to provide telestroke support to their referral hospitals. Average door-to-needle times for tPA administration for the prior year, across all sites, was 44.4 minutes (range 25-60 minutes), and overall average door-to-groin puncture for patients undergoing EVT was 103.4 minutes (45-150 minutes). At the time of the survey, 5 (55.6%) of these centers reported utilizing a combination of clinical presentation, non-contrasted head CT ASPECTS (Alberta Stroke Program Early CT Score) and CT Angiogram for identifying patients appropriate for EVT. Three centers also utilized CT Perfusion imaging and one center indicated use of MRI/MRA +/- MR Perfusion imaging for this purpose. During EVT procedures, 2 of the responding centers (22.2%) reported using only conscious sedation, while the remaining 7 reported using either conscious sedation or general anesthesia at the discretion of the treating interventionalist and anesthesia provider. The large majority, 87.5%, reported 24/7 in-house anesthesia teams present for these procedures if needed, with one center reporting anesthesia teams are called in on off-hours or weekends.

Admission level of care following tPA administration was split, with 55.6% admitting to a stroke floor or step-down unit, and 44.4% admitting primarily to an ICU setting. For patients treated with EVT, only 22% report admission to stroke floor or step-down unit, with the remaining 77.8% admitted to an ICU. Once admitted, tPA treated patients are primarily managed by Vascular Neurologists at 8 sites, with the remaining site utilizing medicine hospitalists for management. Similarly after EVT, vascular neurologists primarily manage these patients at 7 sites, while hospitalists do so at 1, and vascular neurosurgeons at 1 site.

### Referral centers and transfer processes

The majority (7, 87.5%) reported working with their referral hospitals to develop reperfusion and transfer plans for LVO patients. All centers report having an established process for expedited transfer of stroke patients in need of neurointerventional therapies from their referral hospitals. Only 2 (22%) of responding progressive stroke centers indicated that all of their referral hospitals have 24/7 acute vascular imaging capability, while 6 (66.7%) reported some of their referral centers have this capability, and 1 center indicated none of their referral centers had this capability. Only a single center reported that vascular imaging proof of LVO is required prior to transfer-in from their referral centers, with the remaining indicating clinical suspicion alone was sufficient or LVO proof was preferred but not required prior to transfer. For patients transferred in specifically for potential EVT, the majority of responding centers (5) indicated any transportation method was sufficient, while the remaining 4 allowed only ACLS/critical care transport by air or ground. On arrival to the hub hospital for transferred patients with planned EVT, 5 (55.6%) of responding progressive centers indicated these patients bypass the ED and are transported directly to the neurointervention suite when staff is available immediately.

### Feedback

The majority (66.7%) report providing regular scheduled feedback on treatment times (including EVT, reperfusion success and/or outcomes) to EMS providers; 1 within 24-48 hours, 1 within 7 days and 4 in >7 days. All responding centers have multidisciplinary stroke committees which routinely meet to evaluate stroke performance metrics and make improvement recommendations. 7 centers indicated meeting at least yearly with other hospitals in their region to evaluate performance metrics and make improvement recommendations. Likewise 7 centers report including local EMS agencies corresponding to their referral hospitals during these multidisciplinary meetings. The majority (6, 66.7%) participate in Get With The Guidelines Stroke Data Repository, and an additional two centers participate in an equivalent data repository program.

### Discharge

Only 22% of centers identify patients at high risk for readmission prior to their discharge, and this is based on clinical judgment of the attending physician. All centers reported to have a patient discharge process that is documented in the medical record which includes a focused health care professional interaction including review and reconciliation of discharge medications and patient education materials. Follow up appointments are scheduled at the time of discharge in 77.8% of responding centers, and this follow up occurs within 2 weeks at one center, from 2 weeks to 1 month at one center, and within 90 days in the remaining 5 centers. Only 3 of the responding sites indicate a protocol in place to call patients within 1 week of discharge to check on clinical stability and medication compliance.

## Discussion

We developed this international survey of advanced and progressive stroke centers in order to obtain expert opinion-level data in acute stroke care, and inform advanced practice recommendations for the IMPROVE Stroke Care Program Manual of Operations.

The survey results showed several areas of current controversy are managed variably even in these advanced centers, though trends can be identified. For example, slightly over half of responding centers indicated that transferred-in LVO patients bypass their ED and are transported directly into the neurointerventional suite. Additionally, in large systems, destination protocols to transport patients with suspected LVO directly to thrombectomy-capable centers was routinely practiced in only about half of the responding centers. Only 4 of the 9 responding centers noted the routine use of perfusion imaging modalities for patient selection for intervention; notably, this survey was completed before the release of the DAWN and DEFUSE 3 trial data. Similarly, the level of admission (ICU, step-down unit, or ‘floor’ acute care unit) was split amongst the responding centers, which may indicate the location is less important than the processes and practice in those locations.

Other practices that were expected to be found at some progressive centers were in fact not in use at all, such as the utilization of pre-hospital mobile apps for tracking and communicating stroke patient data. Similarly, some practices that were expected to be universal, were not. The majority, but not all responding centers, gave quick feedback to their respective EMS, ED, and provider teams regarding stroke patient processes and outcomes. The majority, but not all centers, also participate in large-scale data repositories such as Get With The Guidelines or equivalent.

There were several limitations to this study. The response rate was 45%, however as the design of this study was not intended to be a representative sample of hospitals, and was essentially a convenience sampling methodology, it is unlikely this introduced significant bias. Likewise, non-responding centers were fairly evenly distributed across the globe, including 3 European, 1 Canadian and 6 centers in the United States (3 Midwest, 4 ‘stroke belt’/Southeast). More Southeast US sites were invited to participate than other locations. Further, language barrier may have contributed to non-response from some European sites. No Asian sites participated. Another limitation is lack of available outcome data from all participating centers. While this information could be used to more objectively define high-performing centers, it is rarely widely available without pre-existing and time consuming data use agreements. Further, we are not able to verify any responses.

Additionally, some practices varied significantly between North America and Europe. Namely, transportation of patients to specific centers is managed differently in compact and high-density European city centers. For example, in London, all acute stroke patients are transported directly to the advanced centers regardless of suspected LVO status. As another example, in many UK centers, stroke patients are managed by ‘stroke physicians’ who may not be neurology trained and boarded, which affected responses to staffing survey questions.

Overall, this international survey of advanced and progressive stroke systems revealed useful practice patterns and processes, but also highlights that a one-size-fits-all approach is unlikely to be useful. Instead, we suggest regional collaboration to evaluate area resources, geography and hospital capabilities, to develop local best-practices. Processes from other high-functioning stroke systems such as those outlined in our survey should be used as a guide or menu of possible approaches that have a documented history of success in specific settings. The findings of this survey were used to supplement AHA/ASA guideline statements as well as results of big-data and large clinical trial publications in the development of the IMPROVE Stroke Care Manual of Operations recommendations. Practices that were common among the advanced systems surveyed are listed as lower level-of-evidence recommendations for potential changes in practice in local stroke systems when resources or improvement goals align. Controversial practices remain differentially managed by different centers and should be considered in the context of a particular hospital’s resources and overall system of care.

## Supporting information

Appendix 1. Survey Tool

## Data Availability

All data produced in the present study are available upon reasonable request to the authors

