## Appendix 1. Survey Tool for "Current Practices in Stroke Systems of Care: An International Survey of Progressive Stroke System Processes"

### IMPROVE Stroke Progressive Regional Stroke Survey

OBJECTIVE

To understand the elements of progressive regional stroke systems that program leaders utilize to impact stroke patients' outcomes. These best practices will be aggregated and used as a standardized approach to treating stroke in the new era of preferred reperfusion in eligible patients.

Please answer the following questions to the best of your ability.

---

#### CENTER SPECIFIC DEMOGRAPHICS

Name of respondent completing this survey: ________________________________

Please identify your Region name, City, State, and Country: ________________________________

Hospital or academic center that you represent (please include address): ________________________________

How would you classify your stroke center?

- ◯ Stroke Ready Hospital - (i.e. Capable of diagnosing acute stroke, imaging with CT and treating with Alteplase [t-PA])
- ◯ Primary Stroke Center (as defined by the US Joint Commission of Hospital Certification or an equivalent certifying agency)?
- ◯ Intervention Capable Hospital (primary stroke center with endovascular capacity but without credentials of comprehensive stroke center)
- ◯ Comprehensive Stroke Center (as defined by the US Joint Commission of Hospital Certification or an equivalent certifying agency)

#### GENERAL SYSTEM ASSESSMENT

How many ischemic strokes does your center care for in a year? __________________________________

How many primary hemorrhagic strokes (primary intraparenchymal hemorrhages) does your center care for in a year? __________________________________

How many primary subarachnoid hemorrhages (SAH) does your center care for in a year? __________________________________

How many patients are treated with tPA at your center per year? __________________________________

How many endovascular interventions for ischemic stroke are completed at your center in a year? __________________________________

How many aneurysmal SAH patients are treated at your center per year? __________________________________

What perentage of total aneurysmal SAH patients are treated using endovascular methods? __________________________________

What percentage of aneurysmal SAH patients are treated surgically? __________________________________

Do you have a dedicated Stroke Unit?
- ◯ Yes
- ◯ No

Do you have a dedicated Neurocritical Care Unit?
- ◯ Yes
- ◯ No

How many Vascular Neurologists does your center have? __________________________________

How many Vascular Neurosurgeons does your center have? __________________________________

How many Neuro-intensivists does your center have? __________________________________

How many Neuro-interventionalists (these may be from any of the following specialties: radiology, neurosurgery or interventional neurology) does your center have? __________________________________

Is your neurointerventional suite open 24 hrs/day, 7 days/week, 365 days/year?
- ◯ Yes
- ◯ No

Does a neurointerventionalist respond to all stroke codes or evaluate all stroke code imaging to assess eligibility for endovascular therapies?
- ◯ Yes
- ◯ No

Is there a back up interventionalist in case there are multiple cases needing endovascular intervention?
- ◯ Yes
- ◯ No

If your suite(s) or on-call neurointerventionalists are busy, do you have a second neurointerventional center to defer to?
- ◯ Yes
- ◯ No

#### PRE-HOSPITAL SYSTEM ASSESSMENT

Do(es) the EMS agency(s) that support your hospital have training in identifying patients at risk for stroke based on chief complaint or presenting symptoms?

- ◯ Yes - in all vehicles
- ◯ Yes - in >50%, but not all vehicles
- ◯ Yes - in some (< 50%) vehicles
- ◯ No

If yes, do they use an established pre-hospital stroke scale to evaluate the patient?

- ◯ Yes
- ◯ No

If yes, what scale is used? ______________________________

Do you have a system that predetermines EMS destination based on severity of the stroke scale or other factors?

- ◯ Yes
- ◯ No

Have you worked with EMS to develop destination plans for acute stroke patients?

- ◯ Yes
- ◯ No

If yes, what factors beyond patient preference or proximity of center are used to determine destination?

_____________________________________

Does your system use Mobile Stroke Units?

- ◯ Yes
- ◯ No

What type of capabilities for reperfusion do the hospitals have, that EMS in your region take stroke patients to? (please select all that apply)

- ☐ a) Hospitals that do not give Alteplase - Not stroke ready
- ☐ b) Stroke ready (give Alteplase and ship to a primary stroke center)
- ☐ c) Primary Stroke Center, NOT interventional capable
- ☐ d) Primary Stroke Center, neurointerventional
- ☐ e) Neurointerventional, Comprehensive Stroke Center

How many 'not stroke ready' (option 'a' above) hospitals are in your region? ______________________________

How many 'Stroke Ready' (option 'b' above) hospitals are in your region? ______________________________

How many 'Primary Stroke Centers - not interventional capable' (option 'c' above) are in your region? ______________________________

How many 'Primary Stroke Centers - interventional capable' (option 'd' above) are in your region? ______________________________

How many 'Comprehensive Stroke Centers' (option 'e' above) are in your region? ______________________________

Does EMS use commercial technologies (such as PULSARA or any other mobile app) to track stroke time processes?

- ◯ Yes
- ◯ No

If yes, please list app(s): ______________________________

Does your hospital provide regular feedback on time to treatment (including neurointervention times, reperfusion success and/or outcomes) to EMS providers?

- ◯ Yes, within 24-48hrs
- ◯ Yes, within 7 days
- ◯ Yes, in >7 days
- ◯ No

If EMS assessment tools are positive for suspected acute stroke, do patients at your center go directly to imaging (CT, MRI) and bypass evaluation in the Emergency Department?

○ Yes
○ No

If yes, what triggers this Emergency Department bypass?

__________________________________

#### TREATMENT SPECIFIC ASSESSMENT - EMERGENCY DEPARTMENT AND HOSPITAL CARE

What percentage of acute stroke patients in your center are brought to the ED by EMS? ________________________

What percentage of acute stroke patients in your center are brought to the ED by private vehicle (driven by family or friends)? ________________________

Who is responsible for activating "Code Stroke"s at your institution's ED? (select all that apply)
- ☐ Registered Nurse at ED Triage
- ☐ Nurse practitioner / Physician Assistant
- ☐ Medical Doctor

Do you have 24/7 in-house Neurology coverage?
- ◯ Yes
- ◯ No

Do you have Neurologists on staff covering from home?
- ◯ Yes
- ◯ No

Does your center have a dedicated Stroke/Neurovascular service?
- ◯ Yes
- ◯ No

Do you utilize a telestroke service?
- ◯ Yes
- ◯ No

If yes, which of the following best describes the telestroke service your institution utilizes?
- ◯ Commercial national telestroke provider service
- ◯ Commercial regional telestroke provider service
- ◯ Local/Regional academic center providers
- ◯ We use our own providers, and leverage a telestroke system to support our referral facilities

Who makes the final decision on administration of Alteplase (tPA) at your center?
- ◯ Emergency Physician
- ◯ Staff Neurologist (either in-house or form home call)
- ◯ Telestroke Service Neurologist

What is your average door-to-needle time for ischemic stroke patients treated with Alteplase (tPA) over the past year? (please give average time in minutes) ________________________

What is your average door-to-groin puncture time for patients presenting to your emergency department with acute ischemic stroke and undergoing mechanical thrombectomy? (please give time in minutes; average over past year) ________________________

Where are your post-tPA treated patients most typically admitted?
- ◯ Stroke floor or stepdown unit
- ◯ Intensive Care Unit
- ◯ Transferred to a tertiary care center

Where are your post-thrombectomy patients typically admitted:
- ◯ Stroke floor or stepdown unit
- ◯ Intensive Care Unit
- ◯ Transferred to tertiary care center

Who primarily manages post-tPA stroke patients during the acute phase of illness at your center?
- ◯ Hospitalist
- ◯ Vascular Neurologist
- ◯ Vascular Neurosurgeon

Who primarily manages post-thrombectomy stroke patients during the acute phase of illness at your center?

- ◯ Hospitalist
- ◯ Vascular Neurologist
- ◯ Vascular Neurosurgeon
- ◯ Neurointensivist
- ◯ Medical Intensivist (non-neurology trained intensivist)

#### TERTIARY HOSPITAL SPECIFIC ASSESSMENT

Have you worked with your referral hospitals to develop reperfusion and transfer plans for "large vessel occlusion" (LVO) stroke patients?

- ◯ Yes
- ◯ No

Do you offer your referral (non-neurointervention center) hospitals, upon identification of an LVO stroke, "uinversal access" (24/7) activation of your entire stroke team at the hub?

- ◯ Yes
- ◯ No

Do you have a process for the expedited transfer of stroke patients in need of neurointervention from referral hospitals?

- ◯ Yes
- ◯ No

Are your center's referring spoke hospitals capable of and utilize vascular imaging (CTA, MRA, DSA) 24/7 in the evaluation of acute stroke?

- ◯ yes, all referring hospitals have 24/7 vascular imaging capability
- ◯ Only some of our referring hospitals have this capability
- ◯ None of our referring hospitals have this capability 24/7

Does your center require proof of large vessel occlusion (LVO) (via vascular imaging) prior to transfer for thrombectomy?

- ◯ yes, vascular imaging proof of LVO is required prior to transfer
- ◯ no, LVO proof is not required but it is preferred
- ◯ no, clinical suspicion alone is sufficient

For patients transferred from a referral center to a hub center for potential intervention, what transportation methods are utilized?

- ◯ BLS transport
- ◯ Critical Care / ACLS ground transport only
- ◯ Air transport only
- ◯ ACLS ground or Air transport only
- ◯ Any / all of the above

Which of the following are utilized in deciding appropriateness for mechanical thrombectomy?

- ◯ a) Clinical presentation alone
- ◯ b) Clinical presentation plus non-contrast CT - ASPECTS Score
- ◯ c) option (b) plus CT Angiogram
- ◯ d) option (c) plus CT Perfusion imaging
- ◯ e) option (a) plus MRI/MRA, +/- MR Perfusion imaging

If the patient has a confirmed large-vessel occlusion diagnosis at a non-neurointervention center with plan to transfer for neurointervention consult, do they bypass your hub ED and go straight to the Neurointervention suite when staff is available?

- ◯ Yes
- ◯ No

What is the longest time period that you will accept a CT and/or CTA from a referral hospital before repeating imaging prior to providing endovascular therapy at your center?

- ◯ CT/CTA always repeated
- ◯ CT/CTA repeated if study is >60minutes old
- ◯ other

If 'other' above, please elaborate:

__________________________________

During endovascular thrombectomy procedures, what type of anesthesia do your interventionalists typically use?

- ◯ Always general anesthesia with an in-suite anesthesia team
- ◯ Conscious sedation only
- ◯ Either, at the discretion of the treating interventionalist and anesthesia provider if present

For endovascular thrombectomy procedures performed on weekends and/or after hours, does an anesthesia team need to be called in from home?

- ◯ yes
- ◯ no, there are always in house anesthesia providers available for these procedures

Does your hospital provide feedback on time-to-treatment to referral (Transfer non-neurointerventional) hospital providers?

- ◯ Yes, within 48hrs
- ◯ Yes, within 7 days
- ◯ Yes, in >7 days
- ◯ No

#### DISCHARGE PLANNING AND FOLLOW-UP ASSESSMENT

Do you identify patients at high risk for readmission prior to discharge?
- ◯ Yes
- ◯ No

If yes, what risk stratification tool do you use to identify stroke patients at high risk of readmission?
__________________________________

Do you have a patient discharge process that is documented in the medical record and that includes a focused health care professional interaction including review and reconciliation of discharge medications and patient education materials?
- ◯ Yes
- ◯ No

Is a follow-up appointment scheduled at time of discharge?
- ◯ Yes
- ◯ No

If yes, within what time frame is that follow up scheduled to occur?
- ◯ Within 7 days
- ◯ 7 - 14 days
- ◯ 14 - 30 days
- ◯ Within 90 days

On discharge are patients provided information regarding who to call in the event of questions or recurrent symptoms?
- ◯ Yes
- ◯ No

Is a protocol in place at your center to call the patients within 1 week to check on clinical stability and medication compliance?
- ◯ Yes
- ◯ No

Does your center have a dedicated outpatient stroke clinic to which stroke patients are referred on discharge from their acute event?
- ◯ Yes
- ◯ No

**STROKE CENTER OPERATIONS SPECIFIC ASSESSMENT**

| | |
|---|---|
| Do you have a multidisciplinary stroke committee or team that routinely meets (at least monthly) to evaluate your stroke performance and make improvement recommendations? | ◯ Yes<br>◯ No |
| Do you meet (at least yearly) with other hospitals in your region to evaluate stroke performance and make improvement recommendations? | ◯ Yes<br>◯ No |
| Do you include the EMS agencies that correspond to your referral hospitals when you meet? | ◯ Yes<br>◯ No |
| Do you currently participate in Get With The Guidelines Stroke Data Repository or an equivalent? | ◯ Yes, Get With The Guidelines<br>◯ Yes, a different database/repository<br>◯ No |
| What other centralized stroke registry(s) do you participate in for stroke, if any? | __________________________________ |
| Are you currently collecting key process data for the entire system's stroke patient point of entry (EMS, Transfer, Direct admission, Walk-in to the hub center)? | ◯ Yes<br>◯ No |
